# Protocol for a multicentre randomized controlled trial of normobaric versus hyperbaric oxygen therapy for hypoxemic COVID-19 patients

**DOI:** 10.1101/2020.07.15.20154609

**Authors:** Sylvain Boet, Rita Katznelson, Lana A. Castellucci, Dean Fergusson, Michael Gonevski, Hance Clarke, Cole Etherington, Joseph K. Burns, Sylvain Gagné, Daniel I. McIsaac, George Djaiani, Neal W. Pollock, Laurie Proulx, Louise Y. Sun, Kednapa Thavorn, Christopher Wherrett, Calvin Thompson, Stéphane Moffett, Jean Eric Blatteau, Pierre Louge, Rodrigue Pignel, Virginia Roth, Jay R. MacDonald, Monica Taljaard, Ronald A. Booth

## Abstract

**Background:** At least 1 in 6 COVID-19 patients admitted to hospital and receiving supplemental oxygen will die of complications. More than 50% of patients with COVID-19 that receive invasive treatment such as mechanical ventilation will die in hospital. Such impacts overwhelm the limited intensive care unit resources and may lead to further deaths given inadequate access to care. Hyperbaric oxygen therapy (HBOT) is defined as breathing 100% oxygen at a pressure higher than 1.4 atmosphere absolute (ATA). HBOT is safe, including for lungs, when administered by experienced teams and is routinely administrated for a number of approved indications. Preliminary clinical evidence suggests clinical improvement when hypoxemic COVID-19 patients are treated with HBOT.

**Objective:** We aim to determine the effectiveness of HBOT for improving oxygenation, morbidity, and mortality among hypoxemic COVID-19 patients.

**Methods and analysis:** This trial is a sequential Bayesian Parallel-group, individually Randomized, Open, Blinded Endpoint controlled trial. Admitted hypoxemic COVID-19 patients who require supplemental oxygen (without mechanical ventilation) to maintain a satisfying tissue oxygenation will be eligible to participate. The anticipated sample size of 234 patients is informed by data from a treatment trial of COVID patients recently published. The intervention group will receive one HBOT per day at 2.0 ATA for 75 minutes. Daily HBOT will be administered until the patient does not require any oxygen supplementation, requires any type of mechanical ventilation, or until day 10 of treatment. Patients in the control group will receive the current standard of care treatment (no HBOT). The primary outcome of this trial will be the 7-level COVID ordinal outcomes scale assessed on Day 7 post-randomization. Secondary outcomes will include: (a) **clinical** outcomes (length of hospital stay, days with oxygen supplementation, oxygen flow values to obtain a saturation by pulse oximetry ≥90%, intensive care admission and length of stay, days on invasive mechanical ventilation, sleep quality, fatigue, major thrombotic events, the 7-level COVID ordinal outcomes scale on Day 28; mortality, safety); (b) **biological** outcomes (plasma inflammatory markers); and (c) health **system** outcomes (cost of care and cost-effectiveness). Predetermined inclusion/exclusion criteria have been specified. The analytical approach for the primary outcome will use a Bayesian proportional odds ordinal logistic semiparametric model. The primary analysis will be by intention-to-treat. Bayesian posterior probabilities will be calculated every 20 patients to assess accumulating evidence for benefit or harm. A planned subgroup analysis will be performed for pre-specified variables known to impact COVID-19 prognosis and/or HBOT (biologic sex and age).

**Discussion:** Based on the mortality rate and substantial burden of COVID-19 on the healthcare system, it is imperative that solutions be found. HBOT is a non-invasive and low-risk intervention when contraindications are respected. The established safety and relatively low cost of providing HBOT along with its potential to improve the prognosis of severe COVID-19 patients make this intervention worth studying, despite the current limited number of HBOT centres. If this trial finds that HBOT significantly improves outcome and prevents further deterioration leading to critical care for severe COVID-19 patients, practice will change internationally. If no benefit is found from the intervention, then the current standard of care (no HBOT) will be supported by level I evidence.

**Trials Registration:** NCT04500626

## INTRODUCTION

### Background and rationale

Approximately 15 to 20% of COVID-19 patients present with hypoxemic respiratory failure requiring oxygen supplementation.^1^ More than one third of patients that require low flow oxygen on presentation, and 60% of those requiring higher flow oxygen, will proceed to intubation and mechanical ventilation.^2–4^ Intubation and mechanical ventilation in themselves are associated with mortality rates between 50% and 97% in COVID-19 patients.^3–7^ These modalities also overwhelm limited intensive care unit (ICU) resources and may lead to further mortality if patients do not have access to care. Therefore, any intervention that prevents further clinical deterioration and associated invasive treatments for COVID-19 patients may help to sustain the healthcare system.

Clinical hyperbaric oxygen therapy (HBOT) is defined as breathing 100% oxygen pressures >1.4 atmospheres absolute (ATA) in a specialized chamber.^8^ The pathophysiology of severe COVID-19 pneumonia includes decreased oxygen diffusion from the alveoli to the blood, and a massive pro-inflammatory cytokine response.^9^ HBOT is a well-established and safe^10^ method to increase tissue oxygen delivery up to 10-fold at 2 ATA pressure.^11–13^ Hyper-oxygenation of arterial blood with plasma-dissolved oxygen through HBOT has a strong anti-inflammatory^13–16^ effect and may have a direct viricidal effect on SARS-CoV-2. HBOT is currently approved by the Undersea and Hyperbaric Medical Society and regulatory agencies (e.g., Health Canada, Food and Drug Administration), for 14 indications, including both elective (e.g., soft tissue radiation therapy complications, non-healing chronic wounds) and urgent conditions (e.g., carbon monoxide poisoning, decompression sickness, gas embolism).^8,17^

Case series of HBOT treatment in hypoxemic COVID-19 patients from China, France and the United States show clinical benefits, e.g. reduced ICU admission and intubation rates.^18–21^ Given these promising anecdotal results, a multicentre randomized controlled trial (RCT) is warranted to formally evaluate the effectiveness of HBOT for improving oxygenation and reducing morbidity and mortality in this clinical setting. We propose a multicentre RCT involving hypoxemic hospitalized COVID-19 patients to test the hypotheses that HBOT significantly reduces the duration of oxygen, rates of intubation and ICU admission, length of stay and mortality. We hypothesize that HBOT will improve these clinical outcomes and ultimately save lives.

### Objectives

#### Primary objective

To evaluate the effect of HBOT on clinical outcome status in hypoxemic hospitalized COVID-19 patients one week after initiation of HBOT, measured by the 7-level COVID ordinal outcomes scale.

#### Secondary objectives

To evaluate the effect of HBOT on: (a) other clinical outcomes (i.e., length of hospital stay; days with oxygen supplementation; daily oxygen flow values required to obtain saturation values ≥90%; ICU admission; ICU length of stay; days on invasive mechanical ventilation or high flow oxygenation; major arterial and venous thrombotic events, such as stroke, pulmonary embolism, deep vein thrombosis; sleep quality; fatigue; the 7-level COVID ordinal outcomes scale assessed on Day 28, mortality, and safety; (b) biologic inflammatory markers, including markers of immune activation response and inflammation; and (c) cost of care for COVID-19 patients and cost-effectiveness of HBOT.

## METHODS

### Trial design

We will conduct a sequential Bayesian Parallel-group, individually Randomized, Open, Blinded Endpoint (PROBE) controlled trial. We will follow the design approach and statistical analysis plan made publicly available for COVID researchers.^22^

### Study setting

The trial is planned to take place across five centres from three countries: The Ottawa Hospital (Ottawa, Ontario, Canada), Rouge Valley Centenary Hospital (Toronto, Ontario, Canada), and Misericordia Community Hospital (Edmonton, Alberta, Canada).We plan to recruit more centres to join this multicentre RCT in the future and increase the generalizability of the results.

### Ethics and regulatory approval

Prior to patient recruitment, ethical approval will be obtained from Clinical Trials Ontario as well as the local ethics boards for each participating site. Approval will also be obtained from Health Canada for using HBOT to treat COVID-19 patients in Canada and from required regulatory bodies in other counties according to local laws.

### Eligibility criteria

#### Inclusion criteria

Male or non-pregnant female patients, age ≥18 years, that are confirmed COVID-19 positive by RT-PCR or any other validated method; and diagnosed with pneumonia requiring supplemental oxygen to maintain hemoglobin oxygen saturation ≥90% (determined by pulse oximetry [S_p_O_2_]) will be eligible to participate.

#### Exclusion criteria

Children and pregnant women will be excluded from this study. Patients to which any of the following criteria apply will also be excluded from the study: Patient clinical status felt to be incompatible with HBOT, e.g. respiratory failure requiring mechanical ventilation; inability to maintain position during treatment (sitting or semi-recumbent depending on the chamber); inability to effectively understand and communicate with the hyperbaric operator, inability to provide informed consent; inability to spontaneously equalize ears and refusal of myringotomies; hemodynamic instability requiring vasopressors; or contraindications to HBOT (e.g., pneumothorax or respiratory failure requiring mechanical ventilation).

### Intervention

The scheduled HBOT treatment will take place in existing hospital-based hyperbaric chambers (multiplace or monoplace, depending on the site). In the air pressurized multiplace chamber, the patient will sit and breathe via a head tent (hood) filled with oxygen. In the oxygen pressurized monoplace chamber, the patient will be placed in a semi-recumbent position without the need for a hood. The sessions will be supervised by a hyperbaric physician and chamber controller located outside the chamber. The HBOT session will start with compression over approximately 10 minutes while breathing oxygen, until a pressure of 2.0 ATA is reached, which is equivalent to the pressure found at 10 metres of seawater (Figure 1). The compression will be followed by breathing oxygen with intermittent air breaks (two cycles of 25 minutes of oxygen breathing and five minutes of air breathing, for a total of 60 minutes at pressure). The subjects will then breathe oxygen while the chamber is decompressed at approximately 1 meter/min to sea level pressure (approximately 1.0 ATA), with the time extended if needed by the patient to equalize their ears. The total duration of the session at pressure (2.0 ATA) will be 55 min and the total duration of the session, including compression and decompression will be at least 75 minutes, given once a day (Figure 1). In case of a complication in the chamber that requires urgent interruption of HBOT, the routine emergency protocols will be applied per existing protocols in the units.

**Figure 1.**
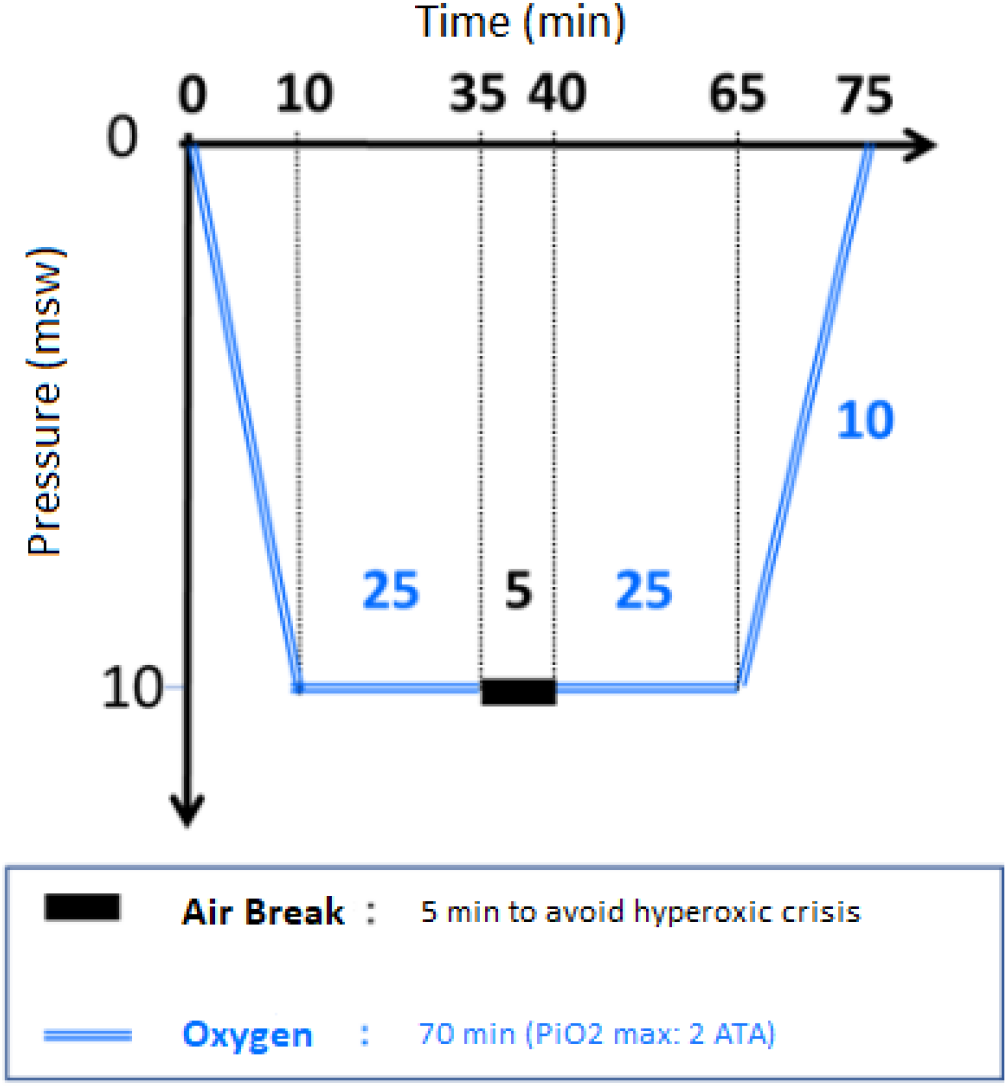
Hyperbaric oxygen therapy profile.

HBOT will be administered on a daily basis until the patient either does not require oxygen supplementation, or deteriorates requiring mechanical ventilation, or up to Day 10. HBOT treatment will be discontinued if a participant misses more than two consecutive days of treatment.

Before the first HBOT session, the hyperbaric operator will educate the patient on wearing the hood and middle ear pressure equalization maneuvers. Before each HBOT session, patients will have their vital signs recorded, contraindications reviewed, and a safety checklist completed. The risks are those inherent to standard HBOT. The hyperbaric units included in this trial are all experienced and conduct several thousands of hyperbaric treatments annually. Compression and decompression speeds are slow to minimize the risk of barotrauma. The risk of oxygen toxicity is minimized with the 2.0 ATA treatment pressure and the five-minute air breaks. The risk of developing an oxygen induced seizure is at most 1/6000 sessions.^10^ In the event the patient complains of respiratory distress during the air break, the air break will be stopped and oxygen breathing resumed.

Between HBOT sessions, patients in the intervention group will receive usual care, including normobaric oxygen supplementation as per standard of care. The control group in this study will receive the current standard of care treatment, including normobaric oxygen.

### Allocation & randomization

The random allocation sequence will be computer-generated by an independent statistician using permuted blocks of randomly varying sizes, stratified by centre, biological sex and age (<60; ≥60 years). COVID-19 epidemiology and prognosis vary by biological sex and age.^3–7^ Study personnel will access the randomization sequence via a central web-based application to ensure allocation concealment.

### Outcomes

Patients will be followed daily during hospitalization and until Day 28, following hospital discharge, or death.

#### Primary Outcome

**The 7-level COVID ordinal outcomes scale assessed on Day 7 post-randomization**, as defined in Table 1. This outcome scale was used in other COVID studies^23^ and was also recommended by the WHO R&D Blueprint expert group.^24^ Ordinal scales have been used as endpoints in clinical trials for other viral pneumonia.^24–27^ We have chosen the ordinal-scaled outcome analyzed with a proportional odds logistic regression model to avoid the loss in statistical power and precision arising from dichotomization.

**Table 1.** Assumed distribution for the primary outcome (COVID ordinal outcomes scale) at Day 7 in the control arm, and in the intervention arm corresponding to an Odds Ratio of 2.

| Outcome code | Control arm<br>(based on NEJM<br>COVID trial)<sup>23</sup> | Intervention arm<br>assuming an Odds<br>Ratio =2 |
| --- | --- | --- |
| 1=death | 7% | 3.6% |
| 2=hospitalized, on ECMO, or requiring invasive mechanical ventilation | 4% | 2.2% |
| 3=hospitalized, on high flow oxygen therapy or non-invasive mechanical ventilation | 21% | 13.2% |
| 4=hospitalized, on supplemental oxygen | 51% | 51.9% |
| 5=hospitalized, not on supplemental oxygen | 17% | 29.1% |
| 6=not hospitalized, unable to resume normal activities | 0% | 0% |
| 7=not hospitalized, able to resume normal activities | 0% | 0% |

#### Secondary Outcomes

**1-Clinical outcomes:** (1) length of hospital stay; (2) days with oxygen supplementation; (3) daily oxygen flow values required to obtain S_p_O_2_ ≥90%; (4) ICU admission, (5) ICU length of stay, (6) days on invasive mechanical ventilation or high flow oxygen; (7) major arterial and venous thrombotic events, e.g., stroke, pulmonary embolism, deep vein thrombosis; (8) self-reported sleep quality; (9) self-reported fatigue; (10) the 7-level COVID ordinal outcomes scale assessed on Day 28; (11) mortality; (12) safety, defined as any adverse events related to HBOT. **2-Biological outcomes:** plasma inflammatory markers (e.g., CRP, D-Dimer). Standard-of-care blood tests results for inflammatory markers ordered for routine care (outside of the study) will be recorded by the research team every day the participant is in hospital. **3-Health system outcomes:** cost of care, as measured by the gold standard client service receipt inventory (CSRI)^28^ at Day 28. The CSRI is adapted to participating countries to capture all social and healthcare care related expenses including financial loss due to health;^28^ Cost-effectiveness, as measured by an incremental cost per one-life year saved.

### Sample size

We estimated the maximum anticipated sample size required using a frequentist approach based on a proportional odds model for the ordinal outcome scale.^29^ Our estimate of 234 patients randomized 1:1 treatment to control is adequate to achieve 80% power to detect an odds ratio (OR) of 2 and assuming the observed distribution of the ordinal scale at Day 7 in the usual care arm of the Cao *et al*., New England Journal of Medicine COVID trial (see Table 1).^23^ The anticipated distribution of the ordinal scale at Day 7 in the intervention arm corresponding to an OR of 2 is also presented in Table 1. We consider differences at level 1-3 to be clinically relevant (e.g., a difference from 7% to 3.6% mortality). If information accruing during the trial differs from that expected, the sample size may change. Sample sizes needed to detect a range of ORs using a proportional odds model are presented in Table 2.

**Table 2.** Required sample sizes to detect Odds Ratios ranging from 1.5 to 2.5.

| Power | OR=1.5 | OR=1.75 | **OR=2** | OR=2.25 | OR=2.5 |
| --- | --- | --- | --- | --- | --- |
| 90% | 920 | 480 | 313 | 229 | 179 |
| 85% | 786 | 410 | 267 | 195 | 153 |
| **80%** | 687 | 359 | **234** | 171 | 134 |

### Data Analysis

#### Primary outcome

Our primary analytical approach will use a Bayesian proportional odds ordinal logistic semiparametric model^30^ with prior distributions for the unknown parameters (Dirichlet distribution for the intercept and skeptical priors for treatment effect as recommended). The summary treatment effect is an OR with OR >1 being favorable to HBOT. The proportional odds model will be adjusted for the following covariates: age; sex; the ordinal outcome scale measured at baseline; time from symptom onset in days; and baseline factors known to be associated with poor outcomes (e.g. diabetes, cardiovascular disease, immunocompromised, long-term care resident). Posterior probabilities will be computed every 20 patients who have completed follow-up. The following posterior probabilities (to be agreed by the trial steering committee and data safety monitoring board [DSMB]) are presented in Table 3 and will be reported on all data accumulated to that point.

**Table 3.**
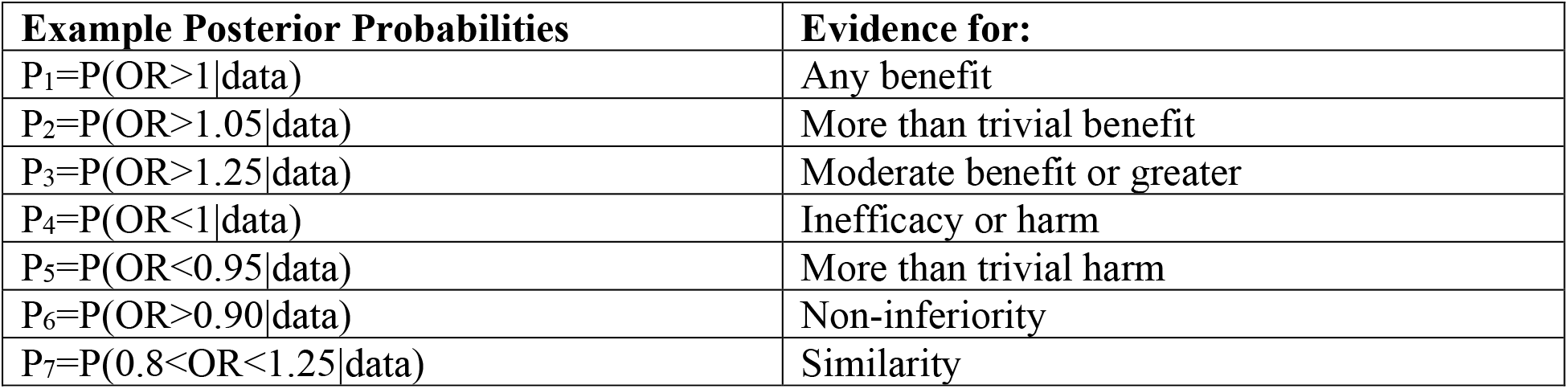
Posterior Probabilities to be reported from the trial.

#### Secondary outcomes

Length of hospital and ICU stay will be analyzed using multistate survival analysis with death modelled as a competing risk. Number of days of oxygen supplementation and days of invasive mechanical ventilation will be analyzed using Poisson or negative binomial regression. Incidence of major thrombotic events, mortality and adverse events related to HBOT will be analyzed using logistic regression or Fisher exact test. Patient-reported outcomes (measures of fatigue and sleep quality) will be analyzed using linear regression. The 7-level COVID ordinal outcomes scale assessed on Day 28 will be analyzed similarly to our primary outcome; biological outcomes will be analyzed using repeated measures generalized linear regression. Since evidence on COVID-19 evolves quickly, additional variables and interactions will be considered based on newer evidence published by the end of our trial.

We will conduct a health economic evaluation alongside the trial to compare total costs and health outcomes of HBOT and usual care groups. Costs and outcomes will be assessed within the follow up period of the trial, i.e. 28 days. Data on services used and the clinical benefit of HBOT will be obtained from the concurrent trial. Unit costs for all resources will be obtained from Canadian sources. All costs will be calculated from a perspective of Canada’s publicly health care system and expressed in 2020 Canadian Dollars. The statistical analysis will be conducted in accordance with current guidelines for cost-effectiveness analysis alongside RCTs.^31^ The incremental cost and incremental outcome will be estimated using generalized estimating equations that explicitly allow for the modelling of normal and non-normal distributional forms of repeated measure data.^32^ An incremental cost-effectiveness ratios (ICER) will be estimated as incremental cost per one-life year saved. The ICER will be obtained through the difference in the mean costs of the two strategies divided by the difference in the mean value of each clinical outcome as denoted by the coefficient of HBOT variable. Uncertainty in the analysis will be addressed by estimating 95% confidence intervals using a non-parametric bootstrapping method. For this study, we will obtain 5000 estimates of costs and outcomes for each strategy. Results from the bootstrapping exercise will also be used to create cost-effectiveness acceptability curves, which represents the probability of HBOT being cost-effective over a range of potential threshold values that the health system may be willing to pay for an additional unit of effect.^33^

#### Planned subgroup analyses

The primary and secondary outcomes will be analyzed for effect modification (treatment X subgroup interaction terms) by pre-specified factors that are known to have an impact on COVID-19 prognosis and/or HBOT: biological sex and age.^3–7^

#### Interim analyses

Safety and study-related adverse events will be reviewed regularly by an independent DSMB. The DSMB will meet every 20 patients that have completed follow-up. The DSMB will call for study suspension and review if they find that one strategy is clearly harmful or beneficial using rules agreed upon by the Trial Steering Committee and DSMB, for example, with evidence for efficacy of P_1_>0.95, with evidence for moderate or greater efficacy if P_2_>0.8, with evidence for inefficacy if P_4_>0.8, or with evidence for harm if P_5_>0.75.

### Trial management

The trial’s coordinating centre will be the Ottawa Hospital Research Institute. The Trial Steering Committee will oversee trial management and will meet as needed. The DSMB will operate independently but will act in an advisory role to the Trial Steering Committee.

### Adverse events

Reporting and handling adverse events will be in accordance with the established protocols at each participating centre. The potential risks that may be encountered by participants in this study are those inherent in a standard HBOT. The side effects or complications are ether minor, reversible, or extremely rare: middle ear discomfort, pain or barotrauma (43%), dental pain (3%), progressive myopia (25%), pulmonary barotrauma (< 0.001%), seizures (0.01%), and hypoglycemia in diabetic patients (1%).^10^

## DISCUSSION

The current SARS-CoV-2 viral pandemic has infected nearly 200 million individuals worldwide and linked to over 4,000,000 deaths as of July 27, 2021.^34^ Based on the case fatality rate and substantial burden of COVID-19 on the healthcare system, it is imperative that a solution be found. HBOT is a non-invasive and low-risk intervention when contraindications are respected, including for lungs.^10,35^ HBOT is also very low risk for healthcare professionals who supervise the treatment. HBOT costs between C$120 and C$500 per treatment depending on the type of chambers and personnel costs.^36,37^ This is significantly less expensive than the daily costs of mechanical ventilation, which is over $3,500 in Canada.^38^ The relatively low cost of providing HBOT along with its potential to improve the prognosis of severe COVID-19 patients make this intervention worth studying, despite the currently limited number of HBOT centres. Evidence of safety, effectiveness, and cost savings would encourage governments to increase access to HBOT in the immediate future and reduce the strain of COVID-19 on the healthcare system. This could be feasibly and quickly accomplished given the availability of medical grade portable chambers (up to 3.0 ATA) and the short time needed to train personnel (within weeks). Increasing access to HBOT may also benefit more patients beyond the current pandemic for already approved indications. If this trial finds that HBOT improves outcome and prevents further deterioration leading to critical care for severe COVID-19 patients, practice will change internationally. If no benefit is found from the intervention, then the current standard of care (no HBOT) will be supported by level I evidence.

## Data Availability

This is a study protocol. No data is available at this time.

## AUTHORS’ CONTRIBUTIONS

Authorship for this trial will be based on involvement in trial design, recruitment, data collection, statistical plan and data analysis. SB drafted the initial protocol. SB, RK, LAC, DF, MT were responsible for conceptualizing trial design. SB and LP managed patient safety protocol. JKB, NE, RK, MG, RP, FS will be responsible for recruitment, enrolment and data collection with the assistance of HC, GD, PL, SG, SM, CT, CW. MT is responsible for power calculation, statistical plan and data analysis. KT is responsible for the cost data collection and cost-effectiveness analysis. VR is responsible for providing operational support. All authors have critically revised the study protocol, with specific details on their areas of expertise, and approved the final version. All authors agree to be accountable for the accuracy and integrity of all aspects of this trial.

## ACKNOWLEDGEMENTS

The authors thank Dr. Felix Soibelman for his help with revising the manuscript and his commitment to future support of this study. The authors thank Sherri Ferguson and Yan Yu He for their help with the regulatory applications.

